# A Randomized Phase II Trial of Gemcitabine, Nab-Paclitaxel, Cisplatin with or without a Medically Supervised Ketogenic Diet for Patients with Metastatic Pancreatic Cancer

**DOI:** 10.1101/2025.06.01.25328728

**Authors:** Gayle S. Jameson, Denise J. Roe, Erkut Borazanci, Diana L. Hanna, Caroline G.P. Roberts, Meredith S. Pelster, Richard C. Frank, Angela T. Alistar, Alan M. Miller, J. Erin Wiedmeier-Nutor, Sandra D. Algaze, Alison R. Zoller, Sarah J. Hallberg, Betsy C. Wertheim, Derek Cridebring, Joshua D. Rabinowitz, Stephen Gately, Jennifer Keppler, Sunil Sharma, Daniel D. Von Hoff, Drew W. Rasco

**Author notes:** **Corresponding Author:** Diana Hanna, USC Norris Comprehensive Cancer Center Los Angeles, CA, USA. Deceased (Sarah J. Hallberg). These authors contributed equally to this work (Gayle S. Jameson, Denise J. Roe, Erkut Borazanci, Diana L. Hanna, Daniel D. Von Hoff, Drew W. Rasco).

## Abstract

**Background:** In this Phase II randomized clinical trial, we evaluated a medically supervised ketogenic diet (MSKD) compared to a usual diet (non-MSKD) when combined with the triplet chemotherapy regimen of gemcitabine, nab-paclitaxel with cisplatin in patients with treatment-naive advanced pancreatic ductal adenocarcinoma (PDAC).

**Methods:** Patients with treatment-naive metastatic PDAC were randomized 1:1 to MSKD or non-MSKD while receiving gemcitabine, nab-paclitaxel, cisplatin on Days 1, 8 of a 21-day cycle. The MSKD was guided by tracking of daily ketone (beta-hydroxybutyrate, BHB) and glucose levels, a web-based application, education, and communication with a remote care team to maintain nutritional ketosis, targeting BHB 0.5–3.0mM. Patients with BMI < 18 kg/m^2^, type 1 diabetes or history of diabetic ketoacidosis were excluded. The primary endpoint was progression-free survival (PFS), tested using a one-sided alpha level of 0.20. Secondary endpoints included overall survival (OS), disease control rate (DCR; partial response + complete response + stable disease at 9 weeks), incidence and severity of adverse events (AEs) and changes in CA 19-9, fasting insulin, HbA1c, BHB, body weight, and quality of life (QLQ-C30).

**Findings:** Fifty-six patients with untreated metastatic PDAC were consented, of which 41 were eligible and 36 were enrolled and randomized. Among 32 evaluable patients (median age 65.9 years; 53% male), 16 were randomized to each arm. In the MSKD arm, 15 of 16 patients achieved nutritional ketosis at any point during the study, with mean BHB of 0.49 mM (95% CI 0.36–0.63) and median proportion of days in ketosis of 39.4% (range 0-95.8%). The study met its primary endpoint. Patients on the MSKD had a PFS by RECIST or clinical progression of 8.5 months, compared to non-MSKD of 5.5 months, HR (95% CI) = 0.53 (0.20 - 1.36) p = 0.092 (one-sided). Patients in the MSKD arm had a median OS of 13.7 months versus 10.2 months in the non-MSKD arm, HR (95% CI) = 0.58 p = 0.107 (one-sided). All MSKD-related AEs were Grade 1-2 and included fatigue, constipation, weight loss, decreased appetite, dehydration, dizziness and nausea. None of the patients stopped the MSKD due to related AEs. There were no significant differences in grade ≥3 chemotherapy-related AEs between the arms.

**Conclusions:** A medically supervised ketogenic diet is feasible in patients with treatment-naïve metastatic pancreatic adenocarcinoma, and when combined with gemcitabine, nab-paclitaxel, and cisplatin, demonstrates significant improvements in progression-free and overall survival, without added toxicity or detriment to quality of life. Larger studies are required to definitively establish the value of ketogenic diet in pancreatic cancer treatment.

## INTRODUCTION

The intersection of diet and cancer therapy has long intrigued clinicians and researchers, with dietary interventions being explored as adjunctive strategies to conventional cancer treatments [1]. Cancer cells exhibit altered metabolism, such as the Warburg effect, where they avidly consume glucose and rely on glycolysis, even in the presence of oxygen [2, 3]. The ketogenic diet, a higher-fat, lower-carbohydrate, moderate-protein dietary regimen, has garnered attention for its potential to exploit the metabolic vulnerabilities of cancer cells [4–9]. By reducing carbohydrate intake, the ketogenic diet results in lower glucose and insulin levels and elevated levels of ketone bodies β-hydroxybutyrate (BHB), acetoacetate, and acetone. BHB is synthesized from circulating fats during fasting and carbohydrate restriction, and serves as an energy carrier, carbon source, reducing agent, and signaling metabolite affecting gene expression, lipid metabolism, and metabolic rate [10].

Pancreatic ductal adenocarcinoma (PDAC) is the third leading cause of cancer-related death in the United States in 2024, and more than 80% of patients present with locally advanced unresectable or metastatic disease [11]. Current treatment regimens for patients with PDAC offer modest improvements in overall survival (OS). A Phase Ib/II prospective study in patients with metastatic PDAC showed promising results evaluating efficacy and safety of triplet therapy (nab-paclitaxel, gemcitabine, cisplatin) in patients with previously untreated metastatic PDAC [12, 13]. However, further improvements to this regimen are needed.

PDAC tumors have profound stromal fibrosis and hypoxia, as well as lower glucose and higher lactate levels compared to surrounding pancreatic tissues, reflecting avid tumor glucose demand [14, 15]. In murine KPC pancreatic cancer models, the combination of a ketogenic diet and triplet therapy synergistically suppressed tumor growth, tripling the survival benefit of chemotherapy alone [16].

Based on these promising preclinical results, we conducted an open label randomized Phase II trial comparing a medically supervised ketogenic diet (MSKD) to a usual diet (non-MSKD) in combination with triplet chemotherapy in patients with treatment-naïve PDAC. Patients randomized to MSKD were actively monitored using a continuous care intervention (CCI) developed and delivered via telemedicine by Virta Health [17, 18]. The CCI includes the use of home biomarker tracking tools, a fingerstick blood glucose and ketone (BHB) meter, a web-based software application for remote data collection, as well as education and communication with a remote care team to actively monitor the development and maintenance of nutritional ketosis. To our knowledge, this is the first randomized controlled trial to utilize a CCI to actively monitor and coach patients to maintain nutritional ketosis, and to evaluate the utility of this nutritional intervention combined with chemotherapy to improve outcomes in patients with PDAC.

## RESULTS

### Patient Characteristics

Among 32 evaluable patients (median age 65.9 years; 53% male), 16 were randomized to each arm. Baseline characteristics were generally well balanced between the arms. Both arms had similar baseline weight, HgbA1c, insulin and BHB levels. CA 19-9 levels and neutrophil-to-lymphocyte ratio were non-significantly higher in the non-MSKD arm at baseline. (Table 1)

**Table 1.** Baseline Patient Characteristics.

| **Characteristic<sup>1</sup>** | **Non-MSKD**<br>***n* = 16** | **MSKD**<br>***n* = 16** | **Total**<br>***n* = 32** | ***P***<br>**value<sup>2</sup>** |
| --- | --- | --- | --- | --- |
| Age (y) |  |  |  |  |
| Median | 65.9 | 66.1 | 65.9 | 0.985 |
| Range | 46.0–75.6 | 40.7–75.8 | 40.7–75.8 |  |
| Weight (kg) |  |  |  |  |
| Median | 76.3 | 73.2 | 74.2 | 0.731 |
| Range | 52.5–120.5 | 45.9–127.0 | 45.9–127.0 |  |
| Glucose<sup>3</sup> (mg/dL) |  |  |  |  |
| Median | 112.5 | 105.5 | 110.5 | 0.485 |
| Range | 84.0–171.0 | 85.0–208.0 | 84.0–208.0 |  |
| HbA1c (%) |  |  |  |  |
| Median | 6 | 5.9 | 6 | 0.545 |
| Range | 5.4– 7.6 | 4.6– 9.0 | 4.6– 9.0 |  |
| Insulin (pmol/L) |  |  |  |  |
| Median | 90.3 | 76.4 | 78.5 | 0.744 |
| Range | 27.8–659.8 | 29.2–138.9 | 27.8–659.8 |  |
| BHB (mmol/L) |  |  |  |  |
| Median | .29 | .26 | .26 | 0.940 |
| Range | 0.08–0.97 | 0.07–1.10 | 0.07–1.10 |  |
| Neutrophil-to-lymphocyte ratio |  |  |  |  |
| Median | 4.27 | 2.55 | 3.35 | 0.119 |
| Range | 1.90–6.88 | 1.65–7.13 | 1.65–7.13 |  |
| Sex, *n* (%) |  |  |  | 0.723 |
| Male | 9 (56.2) | 8 (50.0) | 17 (53.1) |  |
| Female | 7 (43.8) | 8 (50.0) | 15 (46.9) |  |
| Race/ethnicity, *n* (%) |  |  |  | 0.451 |
| Non-Hispanic white | 10 (66.7) | 14 (87.5) | 24 (77.4) |  |
| Hispanic | 3 (20.0) | 1 (6.2) | 4 (12.9) |  |
| Black | 1 (6.7) | 1 (6.2) | 2 (6.5) |  |
| American Indian or Alaska Native | 1 (6.7) | 0 (0.0) | 1 (3.2) |  |
| Karnofsky performance status, *n* (%) |  |  |  | 0.319 |
| 80 | 8 (50.0) | 4 (25.0) | 12 (37.5) |  |
| 90 | 6 (37.5) | 8 (50.0) | 14 (43.8) |  |
| 100 | 2 (12.5) | 4 (25.0) | 6 (18.8) |  |
| History of diabetes, *n* (%) | 5 (31.2) | 4 (25.0) | 9 (28.1) | 0.694 |
| Insulin use, *n* (%) | 3 (18.8) | 3 (18.8) | 6 (18.8) | 1.000 |
| CA 19-9 expressor (>35), *n* (%) | 16 (100.0) | 13 (81.2) | 29 (90.6) | 0.069 |
| CA 19-9 among expressors (U/mL) |  |  |  |  |
| Median | 8954 | 3479 | 5484 | 0.329 |
| Range | 512–336300 | 102–200000 | 102–336300 |  |

<sup>1</sup> Missing data: insulin (*n*=10), BHB (*n*=3), race/ethnicity (*n*=1)<sup>2</sup> Rank-sum test (continuous variables) or chi-square test (categorical variables)<sup>3</sup> Glucose levels were obtained in the clinic and were non-fasting.

Patients on the MSKD arm received a median duration of 8.0 cycles of therapy (range 2.0-19.0 cycles), while the non-MSKD arm had a median treatment duration of 6.5 cycles (range 1.0–16.5 cycles). There was no significant difference in treatment duration between the arms (p = 0.249). Most patients in both arms completed at least 3 cycles of therapy (88% non-MSKD, 94% MSKD).

### Feasibility and Safety of the Medically Supervised Ketogenic Diet (MSKD)

In the MSKD arm, BHB levels were monitored by daily fingerstick tracking which were submitted remotely to the Virta Health team. On days with multiple measurements, the maximum value was used for determining whether the participant was in ketosis.

In both arms, BHB levels were also measured at each site through the institutional laboratory: on the first day of every three cycles in the MSKD arm and on the first day of every cycle in the non-MSKD arm.

Using daily ketone tracking, 15 of 16 patients in the MSKD arm achieved nutritional ketosis (0.5–3.0mM) at any point during the study, with mean BHB levels of 0.49 mM (95% CI 0.36– 0.63) and median proportion of days in ketosis of 39.4% (range 0-95.8%). The mean percentage of treatment days that a participant submitted a BHB measurement was 58.5% (range 3.5-100%). In an exploratory analysis, using a BHB cutoff of ≥ 0.3 mM, all 16 patients achieved ketosis at any point in the study. In contrast, only 6 of 16 patients in the non-MSKD arm achieved ketosis at any point in the study using laboratory data.

Based on laboratory data obtained on the first day of every three cycles, the MSKD arm had a significantly higher change in median BHB levels from C1D1 to C4D1 (median, IQR) (0.04, -0.03 to 0.78) versus the non-MSKD arm (– 0.05, -0.41 to -0.02) arm (p = 0.021).

All MSKD-related adverse events (AEs) were Grade 1-2 and included fatigue (n=3), constipation (n=3), weight loss (n=3), decreased appetite (n=1), dehydration (n=1), dizziness (n=1), nausea (n=1) and body ache (n=1). The incidence of these Grade 1-2 MSKD-related AEs was not different than those reported by MSKD participants that were unrelated to diet, or the incidence in non-MSKD participants. None of the patients stopped the MSKD due to AEs.

There were no unexpected chemotherapy-related AEs and no treatment-related deaths on study. (Table 2, Supplementary Table S1, and <u>Table S2</u>)

**Table 2.** Summary of Grade ≥ 3 Chemotherapy-Related Adverse Events.

| Adverse Event | Non-MSKD<br>(n = 16) | MSKD<br>(n = 16) |
| --- | --- | --- |
| Anemia | 9 (56.3%) | 6 (37.5%) |
| Neutrophil Count Decreased | 6 (37.5%) | 8 (50%) |
| Febrile Neutropenia | 1 (6.2%) | 0 |
| Platelet Count Decreased | 13 (81.2%) | 9 (56.2%) |
| WBC Decreased | 2 (12.5%) | 2 (12.5%) |
| Dehydration | 0 | 1 (6.2%) |
| Hypokalemia | 2 (12.5%) | 1 (6.2%) |
| Hyponatremia | 1 (6.2%) | 0 |
| Hypophosphatemia | 1 (6.2%) | 0 |
| Peripheral Sensory Neuropathy | 0 | 1 (6.2%) |
| Hematuria | 1 (6.2%) | 0 |
| Pulmonary Embolism | 0 | 1 (6.2%) |
| Sepsis | 1 (6.2%) | 0 |
| Colitis | 1 (6.2%) | 0 |
| Diarrhea | 3 (18.8%) | 0 |
| GI hemorrhage | 1 (6.2%) | 0 |
| Intra-abdominal hemorrhage | 1 (6.2%) | 0 |
| Nausea | 0 | 1 (6.2%) |
| Vomiting | 0 | 1 (6.2%) |
| Aspartate transaminase increased | 0 | 1 (6.2%) |
| Fatigue | 0 | 1 (6.2%) |
| Mucosal Inflammation | 1 (6.2%) | 0 |

### Efficacy and Survival Outcomes

The endpoints of progression-free survival (PFS) and overall survival (OS) were examined using a one-sided alpha level of 0.20. All other endpoints were assessed using a two-sided alpha level of 0.05.

Using RECIST criteria alone, patients on the MSKD arm had a median PFS of 8.5 months, as compared to 6.2 months on the non-MSKD arm, HR 0.70 (95% CI, 0.25-1.96, one-sided p = 0.25). (Table 3) There were 3 patients on the non-MSKD arm who discontinued study therapy for clinical progression without an end-of-treatment imaging response assessment. Based on RECIST or clinical progression, patients in the MSKD arm had a significantly improved PFS compared to the non-MSKD arm, 8.5 versus 5.5 months, HR 0.53 (95% CI, 0.20-1.36, one-sided p = 0.092). (Figure 1)

**Figure 1:**
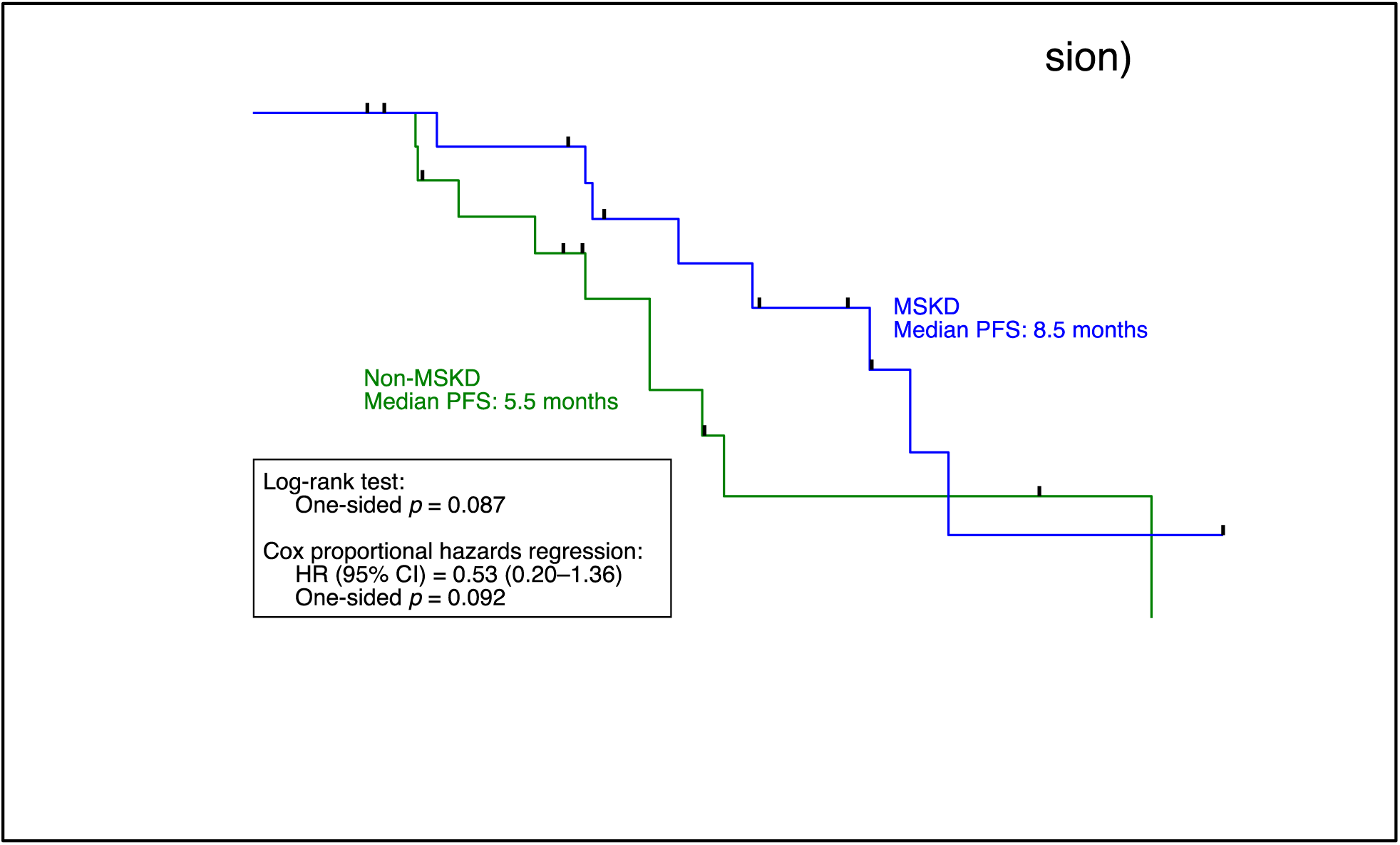
Progression-Free Survival by RECIST or Clinical Progression.

**Table 3.** Progression-Free Survival (PFS), Overall Survival (OS) and Disease Control Rate (DCR)

| Outcome | Non-MSKD<br>(n = 16) | MSKD<br>(n = 16) | *P*<br>two-sided | *P*<br>one-sided |
| --- | --- | --- | --- | --- |
| **PFS (RECIST)** |  |  |  |  |
| Failure, n (%) | 7 (43.8%) | 8 (50.0%) |  |  |
| Median survival (95% CI), months | 6.2 (3.9– .) | 8.5 (4.7–9.6) |  |  |
| Log-rank test |  |  | 0.496 | 0.248 |
| Hazard ratio (95% CI)<sup>1</sup> | 1.0 (ref.) | 0.70 (0.25–1.96) | 0.500 | 0.250 |
| **PFS (RECIST or clinical progression)** |  |  |  |  |
| Failure, n (%) | 10 (62.5%) | 8 (50.0%) |  |  |
| Median survival (95% CI), months | 5.5 (2.8– .) | 8.5 (4.7–9.6) |  |  |
| Log-rank test |  |  | 0.175 | 0.087 |
| Hazard ratio (95% CI)<sup>1</sup> | 1.0 (ref.) | 0.53 (0.20–1.36) | 0.184 | 0.092 |
| **OS** |  |  |  |  |
| Failure, n (%) | 13 (81.2%) | 9 (56.2%) |  |  |
| Median survival (95% CI), months | 10.2 (4.7–<br>16.0) | 13.7 (9.1– .) |  |  |
| Log-rank test |  |  | 0.209 | 0.104 |
| Hazard ratio (95% CI)<sup>1</sup> | 1.0 (ref.) | 0.58 (0.25–1.37) | 0.214 | 0.107 |
| **Best Overall Response** |  |  |  |  |
| Complete Response (CR) | 0 (0.0%) | 0 (0.0%) |  |  |
| Partial Response (PR) | 5 (31.2%) | 11 (68.8%) |  |  |
| Stable Disease (SD) | 9 (56.2%) | 4 (25.0%) |  |  |
| Progressive Disease (PD) | 2 (12.5%) | 1 (6.2%) |  |  |
| DCR at 9 weeks<sup>2</sup> | 14 (87.5%) | 15 (93.8%) |  |  |

<sup>1</sup> Cox proportional hazards regression model<sup>2</sup> Includes two patients whose end-of-study scans occurred at 6.0 and 8.7 weeks, respectively.

Furthermore, patients in the MSKD arm had a significantly improved median OS compared to those on the non-MSKD arm, 13.7 versus 10.2 months, HR 0.58 (95% CI 0.25 – 1.3, one-sided p = 0.107). (Figure 2)

**Figure 2:**
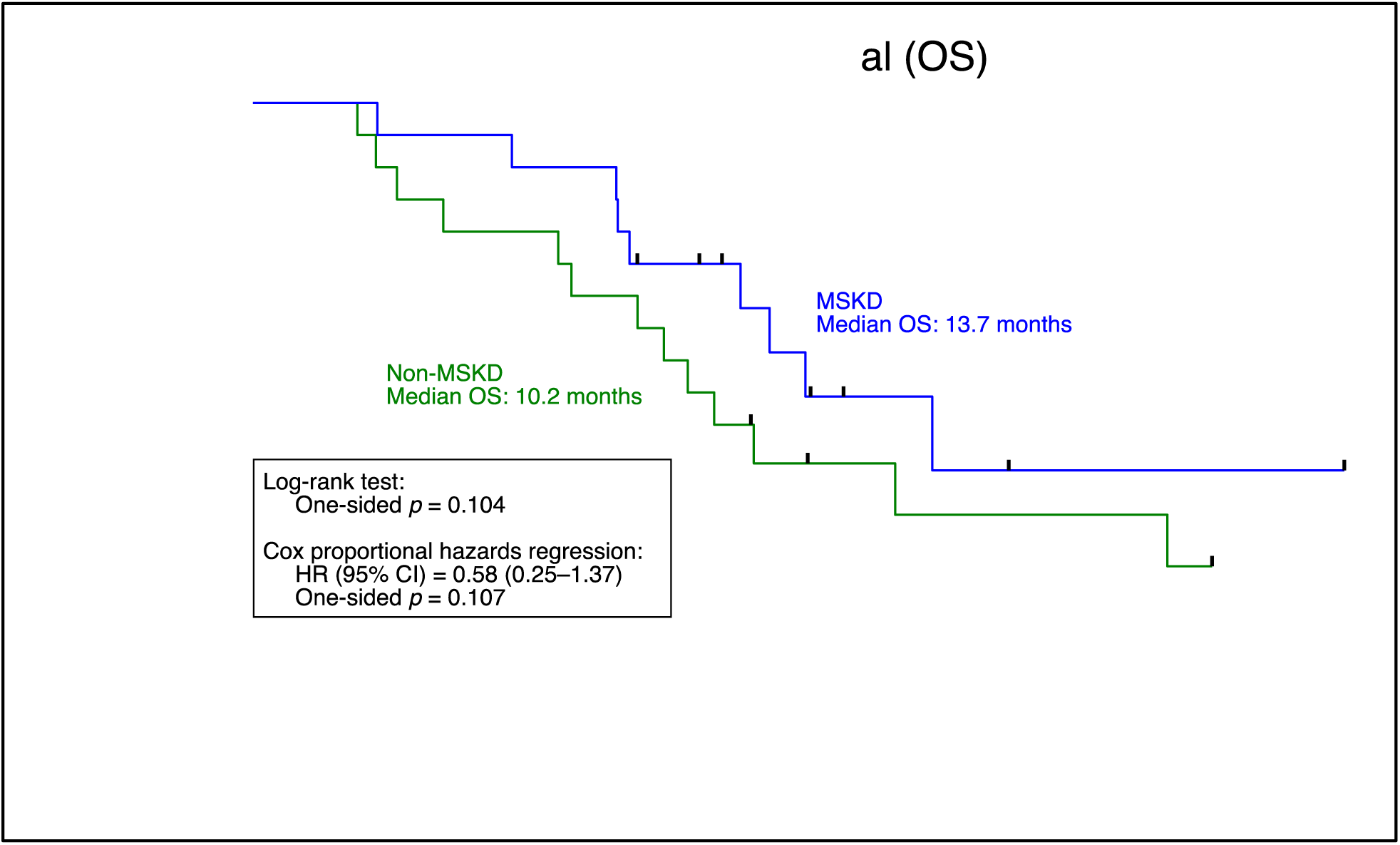
Overall Survival.

The disease control rate (DCR) at 9 weeks did not significantly differ between the non-MSKD (87.5%) and MSKD arms (93.8%) (p = 1.000). More patients on the MSKD arm achieved a best overall response of partial response (PR) compared to the non-MSKD arm, but this did not reach statistical significance (68.8% versus 31.2%, p = 0.110). (Table 3) The maximum change in target lesion tumor burden (an exploratory endpoint) did not significantly differ between the two arms (p = 0.090). (Figure 3) Figure 3 is the swimmers plot illustrating time on treatment, RECIST progression, clinical progression, death, and time at censoring for alive patients.

**Figure 3:**
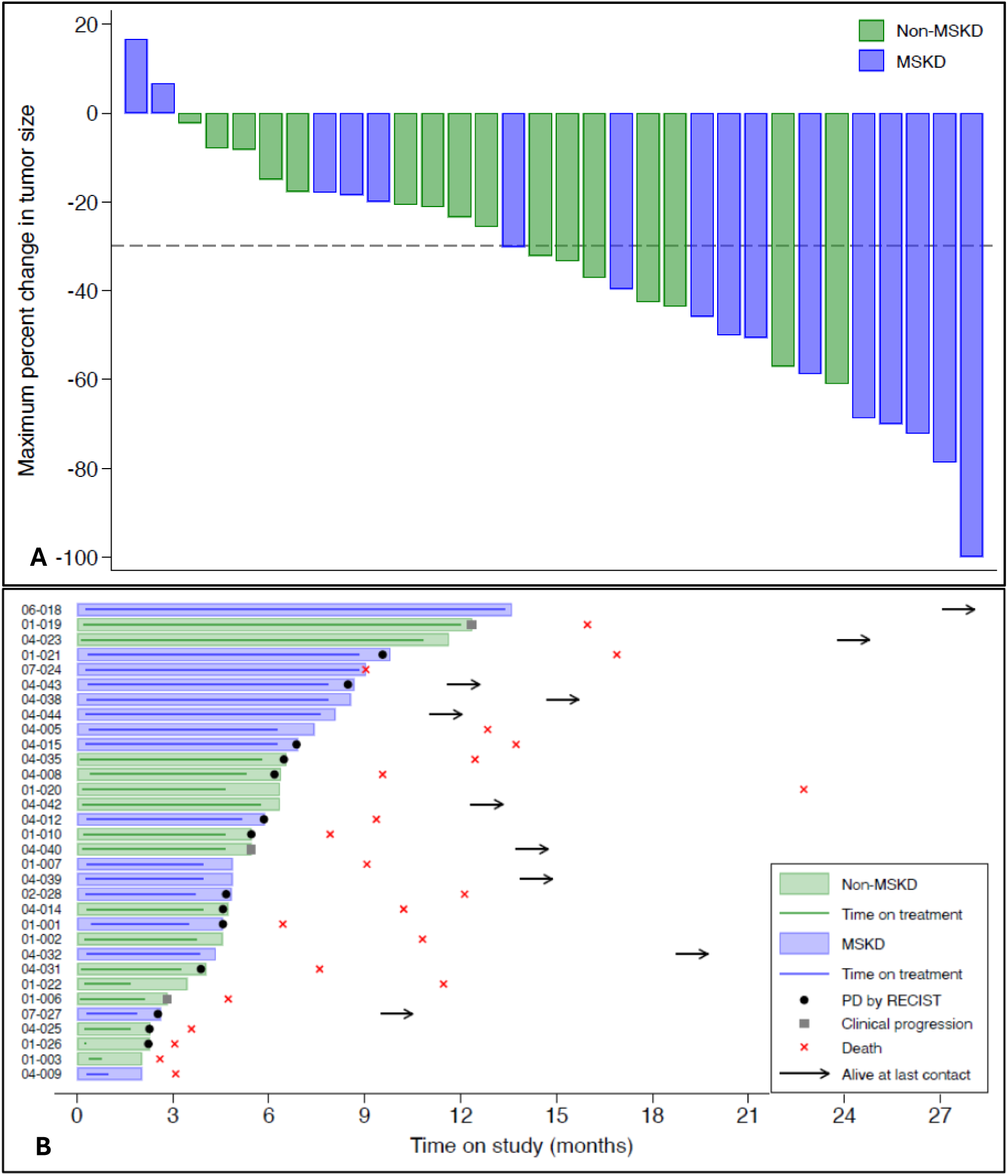
Efficacy Outcomes. **Panel A** shows a waterfall plot of the maximum percent change from baseline in target lesion tumor burden (sum of the longest diameters) per investigator assessment by treatment arm. **Panel B** shows a swimmer plot of outcomes, including time on treatment, PD by RECIST, clinical progression and patient status by treatment arm. Arrows indicate that the patient is alive at last contact.

Patients in the MSKD arm had a greater CA 19-9 decline than those in the non-MSKD arm (linear mixed effect model, p-interaction = 0.076). There was no significant difference in the rate of CA 19-9 normalization between the two arms, MSKD 23% versus non-MSKD 6% (p = 0.299).

### Changes in Metabolic Parameters and Quality of Life Measures

Metabolic parameters were similar between the arms at baseline. (Table 1) Over the course of the study, patients in the non-MSKD arm gained more weight than in the MSKD arm (linear mixed effects model, p-interaction < 0.001). Importantly, there was no significant weight loss and weight remained stable between C1D1 and C4D1 in the MSKD arm (data not shown).

During study treatment, glucose levels were higher in the non-MSKD arm than in the MSKD arm (AUC, p = 0.019). Similarly, glucose levels increased more across time in the non-MSKD arm than in the MSKD arm (linear mixed effects model, p-interaction = 0.072). Furthermore, HbA1c levels decreased more across time in the MSKD arm compared to the non-MSKD arm (linear mixed effects model, p-interaction = 0.076). No significant differences between arms were seen in insulin levels for the study duration.

With respect to QOL measures, EORTC QLQ-C30 Global Health Status and Summary scores did not significantly differ at baseline in both arms. Over the course of therapy, Summary scores increased more in the non-MSKD arm as compared to the MSKD arm (linear mixed effects model, p-interaction = 0.005). In the MSKD arm, Summary scores remained unchanged from C1D1 to C4D1. Similarly, QLQ-C30 Global Health Status scores increased significantly more in the non-MSKD arm between screening and C4D1, compared to the MSKD arm (Wilcoxon rank-sum test, p = 0.045). Importantly, there was no significant decline in either QOL score in the MSKD arm. (Table 4)

**Table 4.** EORTC QLQ-C30 Global Health and Summary Scores During Study Treatment.

| Score | Non-MSKD<br>(n = 15) | MSKD<br>(n = 16) | P |
| --- | --- | --- | --- |
| **Global Health Score** |  |  |  |
| Median (IQR) |  |  |  |
| Screening | 58.3 (41.7, 75.0) | 70.8 (58.3, 91.7) |  |
| C4D1<sup>1</sup> | 75.0 (66.7, 83.3) | 66.7 (50.0, 83.3) |  |
| Change, screening to C4D1<sup>1</sup> | 16.7 (0.0, 25.0) | 0.0 (-8.3, 0.0) | 0.045<sup>2</sup> |
| Maximum change from C1D1 | -12.5 (-33.3, 33.3) | -16.7 (-33.3, 25.0) | 0.553<sup>2</sup> |
| Normalized AUC | 70.8 (52.0, 80.3) | 77.1 (61.0, 79.4) | 0.498<sup>2</sup> |
| Linear mixed effects model | 0.208 (0.120)<sup>3</sup> | -0.013 (0.093)<sup>3</sup> | 0.145<sup>4</sup> |
| **Summary Score** |  |  |  |
| Median (IQR) |  |  |  |
| Screening | 72.6 (60.7, 82.1) | 80.0 (60.2, 93.1) |  |
| C4D1<sup>1</sup> | 84.1 (75.8, 89.7) | 80.0 (66.7, 83.7) |  |
| Change, screening to C4D1<sup>1</sup> | 7.6 (-4.8, 17.1) | 0.1 (-1.4, 3.9) | 0.133<sup>2</sup> |
| Maximum change from C1D1 | 14.0 (-20.4, 17.1) | -10.5 (-19.7, 13.6) | 0.313<sup>2</sup> |
| Normalized AUC | 84.3 (73.6, 89.7) | 85.5 (70.3, 89.2) | 0.918<sup>2</sup> |
| Linear mixed effects model | 0.261 (0.073)<sup>3</sup> | 0.001 (0.057)<sup>3</sup> | 0.005<sup>4</sup> |

<sup>1</sup> Sample size for C4D1: n = 11 (non-keto diet), n = 11 (keto diet)
<sup>2</sup> Wilcoxon rank-sum test
<sup>3</sup> Beta coefficient (standard error) for effect of time on summary score
<sup>4</sup> Test for arm-by-time interaction

## DISCUSSION

This randomized phase II trial is the first to evaluate the impact of a medically supervised ketogenic diet on outcomes in patients with metastatic pancreatic cancer receiving triplet chemotherapy. Our study demonstrated that the MSKD was feasible with an acceptable safety profile and was associated with significantly improved progression-free and overall survival.

Fasting as a precursor to ketosis was described by ancient civilizations[19], and the “ketogenic diet” was first formally described over 100 years ago as an effective therapy for epilepsy[20]. Most evidence assessing the ketogenic diet as an adjunctive cancer therapy lies in glioblastoma[21]. A systematic analysis of glioma studies suggested a survival benefit with the ketogenic diet, but the studies were of limited size[22]. Similarly, ketogenic diet trials in breast and other solid tumors have been small, heterogeneous and shown mixed findings[23–27].

In a recent study of immunocompetent mice implanted with DLBCL tumors, the ketogenic diet significantly increased BHB levels and improved tumor control and overall survival as compared to high-fiber, high-fat high-protein, and high-cholesterol diets[28]. Similarly, in immunodeficient murine xenograft cancer models undergoing CAR-T therapy, BHB supplementation resulted in higher peripheral CAR-T cell expansion, elevated serum effector cytokines and more frequent tumor responses compared to non-BHB-treated controls. These effects were seen in both hematologic and solid tumors, including diffuse large B-cell lymphoma (DLBCL), B-cell ALL (anti-CD19 CAR-T) and pancreatic cancers (anti-mesothelin CAR-T)[28]. In patients with large B-cell lymphoma undergoing CART19 treatment, there was a significant association between BHB serum concentration and CART19 expansion. Furthermore, supplementing CAR-T cells from leukapheresis products with BHB boosted patient T-cell proliferation, with a four-fold increase in absolute cell count[28]. Indeed, accumulating evidence supports BHB as an important anti-tumor effector of the ketogenic diet.

In the preclinical impetus for this trial, adding the ketogenic diet to triplet chemotherapy in murine KPC pancreatic tumor models, led to decreased tumor glucose use, enhanced 3-hydroxybutyrate use and boosted reactive oxygen species, leading to more durable benefits from chemotherapy[16]. Further, glutamine metabolism inhibitors combined with a ketogenic diet had robust effects in these murine models, revealing an additional metabolic vulnerability to be explored[29, 30]. Mechanistically, BHB has far-reaching effects on cancer cell metabolism, epigenetic and transcriptional regulation and the gut microbiome[31]. Importantly, BHB has been shown to serve as a histone deacetylation (HDAC) inhibitor[31]. Furthermore, ketogenic diet-mediated pancreatic tumor regression has been linked to reduction in circulating insulin levels and decrease in the preferred fuel source of glucose which leads to inhibition of ERK and AKT pathways as well as upregulation of JAK/STAT3 and interferon signaling[16, 31, 32].

Diet affects cancer development and recurrence[33], but the role of nutritional interventions as cancer therapy remains elusive and controversial. In a recent systematic review of all randomized controlled trials (RCTs, n=252) examining dietary practices in cancer, only 10% had cancer measurements or survival as endpoints, and adequately powered RCTs failed to demonstrate an improvement in clinical outcomes[34]. Difficulties in translating preclinical and observational findings to the bedside stem from patient, trial, and systems-based factors.

Consistently following a prescribed diet is difficult, and patients with advanced pancreatic cancer have compounding challenges limiting dietary compliance, including taste alterations, abdominal symptoms, anorexia-cachexia, asthenia and depression. While certain patients will harness the opportunity of a medically supervised diet as needed guidance and a part of their disease they can control, others may find the routine monitoring burdensome and the allowed diet restrictive. Importantly, in our study, no patients assigned to the MSKD discontinued the diet due to adverse events, and there were no significant differences in treatment-related AEs, including cytopenias, gastrointestinal effects, or peripheral neuropathy between the arms.

Notably, patients on the non-MSKD arm experienced improved QLQ-C30 Global Health Status scores during the first three cycles of chemotherapy, consistent with previous studies of the triplet regimen[12], while those on the MSKD did not demonstrate improvement. However, baseline QLQ-C30 scores were higher in the MSKD arm, with post-treatment QLQ-C30 scores similar across both arms. Thus, although QLQ-C30 Summary scores significantly increased in the non-MSKD arm as compared to the MSKD arm over the study duration, this may in part reflect differences in pre-treatment baseline status. While these results may suggest challenges in following the MSKD, QOL comparisons in such small sample sizes must be interpreted with caution.

With respect to trial design, most nutritional cancer research to date has been in the preventive or adjuvant settings and reductionist in nature, focusing on one nutrient or food but not addressing the complexity of the whole diet or guiding patients with active cancer on therapy[33, 34]. Conversely, even specific diets retain a degree of heterogeneity, which makes discerning the anti-cancer element complex. Furthermore, dietary interventional studies often rely on patient recollection and assume accurate documentation of intake into diaries which makes capturing compliance cumbersome and less reliable.

The MSKD circumvents these challenges by utilizing a continuous care intervention and relying on goal BHB levels as an objective measure and biologic correlate of compliance. Using a benchmark BHB level of 0.5–3.0mM and with a median 39.4% of days in ketosis, the MSKD led to significant and clinically meaningful improvements in progression-free and overall survival. It is critical to note that ketogenic diets examined in the literature widely vary with respect to how ketosis is defined and achieved, including different caloric allowances and proportions of fats, proteins and carbohydrates. The MSKD in this study allows eating to satiety within the prescribed macronutrient ratios, with the aim of balancing adherence with efficacy.

Merging mechanistic rationale with insights from this initial clinical experience, future investigations should aim to: 1) optimize the level, duration and timing of ketosis alongside systemic therapy; 2) explore the synergy between the MSKD and ketone supplementation, sodium-glucose cotransporter 2 (SGLT-2) inhibitors and/or glutamine metabolism inhibitors; 3) determine the utility of non-invasive markers and metabolites (e.g. insulin, glucose, gut microbiome) in guiding the use of the MSKD; 4) integrate the MSKD into the peri-operative setting and with other treatment modalities (e.g. radiation) and novel therapeutics; 5) examine the benefit of the MSKD in clinically relevant subgroups including those defined by demographic characteristics, tumor gene expression profiles and radiographic correlates (i.e. sarcopenia indices, site of tumor metastases).

### Limitations of the Study

Our study has its strengths and limitations. The primary strength is the randomized design, but the small sample size is a key limitation. Our study used gemcitabine, nab-paclitaxel, and cisplatin; whether the MSKD yields clinical benefit with more commonly used first-line regimens (gemcitabine plus nab-paclitaxel, FOLFIRINOX) is unknown. The frequency and method of BHB measurements varied between the arms, which limits the strength of this comparison. We acknowledge that broad application of the MSKD model may not be practical in certain socioeconomic contexts, including underserved populations or patients without access to smartphones. If larger trials confirm the clinical benefit demonstrated in our study, we would advocate for efforts to expand the reach of ketogenic interventions to all patients with advanced pancreatic cancer.

### Conclusions

The MSKD is feasible and safe and improves progression-free and overall survival in patients with treatment-naïve metastatic pancreatic cancer receiving gemcitabine, nab-paclitaxel and cisplatin. Our trial demonstrates the feasibility of conducting a multi-institutional randomized dietary intervention in a medically complex patient population and paves the way for larger studies which are warranted.

## METHODS

### Ethics and Compliance

The study protocol in the Consort Diagram Supplement 1 and all amendments were approved by the institutional review boards or independent ethics committee at each participating institution. All patients provided written informed consent in a manner consistent with the Declaration of Helsinki [35] prior to participation in the study.

### Trial Registration

This trial is registered under the name ‘Randomized Phase II Trial of Two Different Nutritional Approaches for Patients Receiving Treatment for Their Advanced Pancreatic Cancer (NCT04631445). Trial Pre-Registration: 10/26/2020. Study Start: 11/17/2020.

### Patient Selection

Eligible patients were 18 years or older, had histologically or cytologically confirmed metastatic pancreatic ductal adenocarcinoma, were not previously treated for their metastatic disease that was measurable according to the Response Evaluation Criteria in Solid Tumors (RECIST) version 1.1, [36] had a Karnofsky Performance Status score of 70% or higher, life expectancy of 12 weeks or longer, and adequate hematologic, hepatic, and renal function. Additional eligibility criteria are listed in the trial protocol in the Consort Diagram **Supplement 1**.

Exclusion criterion included: previous radiotherapy, surgery, chemotherapy or investigational therapy for the treatment of their metastatic pancreatic disease (prior treatment in the adjuvant setting with chemotherapy and radiation was allowed, provided at least 6 months had elapsed since completion of the last therapy and recurrence and no lingering toxicities were present); evidence of central nervous system (CNS) metastasis negative imaging study required if clinically indicated, within 4 weeks of Screening Visit; unwillingness or inability to comply with procedures required in this protocol, including unwillingness to follow a ketogenic diet; severe malnutrition or body mass index (BMI) < 18; Albumin < 3.0 g/dL; History of Type 1 diabetes; History of diabetic ketoacidosis (DKA). Additional exclusion criteria are listed in the trial protocol in the Consort Diagram **Supplement 1**.

### Study Design and Treatment

The CONSORT Diagram is shown in <u>Figure S1</u>. Patients randomized to the MSKD arm received remote support, health coaching, daily biometric feedback, and peer support via the Virta Clinic. These patients received nutritional and behavior change educational content prior to starting chemotherapy and continuing for the duration of treatment. Access to a web-based software application was provided to patients in the MSKD arm for biomarker reporting and monitoring, education, and communication with remote care team (via telemedicine) consisting of a health coach and medical provider. Dietary recommendations were to restrict total dietary carbohydrates to ≤30 g per day (typically ≤10% of total energy intake). Daily protein intake was targeted to a level of 1.5 g per kg of reference (ie, medium-frame ideal) body weight and participants were coached to incorporate dietary fats to satiety. Other aspects of the diet were individually prescribed to ensure safety, effectiveness, and satisfaction, including consumption of 3-5 servings of non-starchy vegetables and adequate mineral and fluid intake for the ketogenic state. A meal shipment to cover the patient’s first 24-28 hours on the ketogenic diet was provided. The meals were prepared by a company dedicated to pre-prepared meals meeting specific dietary standards.

Patients in the MSKD arm monitored BHB levels twice daily via a home blood ketone monitor, during the first month, then daily (p.m.) thereafter, and daily (a.m.) glucose monitoring via a home glucometer. These values were uploaded to the Virta-based app for close monitoring and nutritional adjustments as needed by the Virta health coach and/or medical provider. Real-time monitoring by the Virta team allowed individualized nutrition recommendations to achieve and sustain nutritional ketosis with a goal of 0.5-3.0 mmol/L blood BHB. Patients were encouraged to report daily hunger, cravings, energy, and mood. These ratings and BHB concentrations were utilized to adjust nutritional guidance.

Patients randomized to the non-MSKD arm did not have any nutritional intervention provided as part of their participation in the trial and served as the control arm. Patients in this arm met with a local dietician at baseline as per the individual site standard of care. They were instructed to follow their regular diet and not a ketogenic diet. Follow-up with the site-based dietician was per standard of care.

Initial consultation with the Virta Health registered dietitian (RD) for patients in the MSKD arm and with an on-site RD for patients in the non-MSKD arm occurred within 21 days prior to the first dose of chemotherapy.

All patients received intravenous (IV) nab-paclitaxel 125 mg/m^2^, gemcitabine 1000 mg/m^2^, and cisplatin 25 mg/m^2^ on Days 1 and 8 every 21 days until development of unacceptable toxicity or disease progression.

All patients received IV premedication of dexamethasone, 12 mg, palonosetron, 0.25 mg, and fosaprepitant, 150 mg, followed by oral dexamethasone, 4 mg, and ondansetron, 8 mg, twice daily for 2 days after each chemotherapy. The sequence of drug administration was IV hydration followed by nab-paclitaxel, then cisplatin, and then gemcitabine. Patients received additional IV hydration on Days 2 and 9 and received pegfilgrastim, 6 mg, subcutaneously on Day 9 of each cycle as primary prophylaxis.

The study design is given in detail in the Consort Diagram **Supplement 1**.

### Study Assessments

Contrast-enhanced CT evaluations were performed once every 3 cycles (prior to Cycles 4, 7, 10, etc). AEs were assessed according to the Common Terminology Criteria for Adverse Events version 5.0.

### Study Endpoints

The primary trial endpoint was PFS per RECIST 1.1. PFS was defined as the time from randomization to first documentation of objective tumor progression or to death due to any cause.

Secondary endpoints included:

- Overall survival
- The number of complete responses/partial responses as defined by CT scan using the Response Evaluation Criteria In Solid Tumors 1.1 (RECIST 1.1)
- Changes in CA 19-9 (or CA 125, or CEA if non expressers of CA 19-9) and return to normal limits (from at least > 2X ULN).
- Disease control rate using the Response Evaluation Criteria In Solid Tumors 1.1 (RECIST 1.1) - Partial Response + Complete Response + Stable Disease at 9 weeks.
- Change in weight
- Change in insulin levels.
- Change in BHB levels.
- Change in HbgA1c levels.
- Change in quality of life via the European Organization for Research and Treatment of Cancer Quality of Life Questionnaire QLC-C30 (EORTC QLQ-C30) assessment: global health status and summary score.

### Statistical Considerations

This study was designed for a sample size of 32 (16 evaluable patients in each arm), to detect a doubling of PFS from 10 months to 20 months with a power of 80%, assuming a one-sided alpha level of 0.20. The sample size was re-evaluated in July 2023 based on median PFS of 6.2 months observed in the AXCLPANC study[12]. The sample size of 16 evaluable patients in each arm provided > 80% power to detect a doubling of PFS from 6.2 months to 12.4 months (assuming a one-sided alpha level of 0.20). Kaplan-Meier curves were created for each survival outcome, by arm. Differences in survival by arm were tested using log-rank tests and Cox proportional hazards regression. Disease control rate at 9 weeks, best overall response (CR/PR versus other), and CA 19-9 normalization rates were compared using Fisher’s exact tests. Differences in each continuous outcome (CA 19-9, weight, insulin, BHB, HbgA1c, EORTC QLQ-C30 global health status, EORTC QLQ-C30 summary score) were tested in multiple ways: change from screening to C4D1 (Wilcoxon rank-sum test), maximum change from C1D1 (Wilcoxon rank-sum test), normalized AUC (Wilcoxon rank-sum test), and a linear effects model that included an arm-by-time interaction term. Only the primary survival endpoints were assessed using a one-sided alpha level of 0.20; all other endpoints were assessed using a two-sided alpha level of 0.05, and no adjustments were made for multiple comparisons. All statistical tests were conducted using Stata 18 (StataCorp, College Station, TX).

## Data Availability

All data produced in the present study are available upon reasonable request to the authors.

## Acknowledgements

The investigators thank the patients and their families for their participation in this study. We honor the memory of Dr. Sarah J. Hallberg, whose contributions were invaluable to this research.

## Author Contributions

Protocol Design and Writing: G. J., D. R., E. B., D. H., C.R., A. Z., S. H., B. W., D. C., J. R., S. G., D. V. H.

Patient Enrollment: G. J., E. B., D. H., M. P., R. F., A. A., S.A., S. S., D. R.

Meeting Attendance: G. J., D. R., E. B., D. H., C.R., M. P., A. M., A. Z., B. W., D. V. H.

Statistical Analysis D. R., B. W. Dietary Guidance: C. R., A. Z.,

Manuscript Writing: G. J., D. R., E. B., D. H., C.R., M. P., A. A., A. M., A. Z., B. W., D. C., S. G., J. K., D. V. H.

