## Supplemental Data for "A Randomized Phase II Trial of Gemcitabine, Nab-Paclitaxel, Cisplatin with or without a Medically Supervised Ketogenic Diet for Patients with Metastatic Pancreatic Cancer"

[1]HonorHealth Research Institute, Oncology Division, Scottsdale, AZ, USA
[2]The University of Arizona Cancer Center, Cancer Center, Tucson, AZ, USA
[3]USC Norris Comprehensive Cancer Center, Medical Oncology, Los Angeles, CA, USA
[4]Virta Health, Research Department, Denver, CO, USA
[5]Sarah Cannon Research Institute, Nashville, TN, USA
[6]Nuvance Health, Department of Medicine, Norwalk, CT, USA
[7]Atlantic Health, Atlantic Medical Oncology, Morristown Medical Center Morristown, NJ, USA
[8]TD2 Oncology, Scottsdale, AZ, USA
[9] Mayo Clinic, Division of Hematology/Oncology, Phoenix, AZ, USA
[10]Translational Genomics Research Institute (TGen), Clinical Genomics and Therapeutics, Phoenix, AZ, USA
[11]Princeton University, Department of Chemistry, Lewis-Sigler Institute of Integrative Genomics, and Ludwig Institute for Cancer Research, Princeton, NJ, USA
[12]START – San Antonio, San Antonio, TX, USA
[13] Arizona State University, School of Medicine and Advanced Medical Engineering, Tempe, AZ, USA
[14] City of Hope, Department of Medical Oncology and Therapeutic Research, Duarte, CA, USA
[15] Lead Contact

[†]Deceased
[*]These authors contributed equally to this work.

**Corresponding Author:**
Diana Hanna

USC Norris Comprehensive Cancer Center
Los Angeles, CA, USA

### Supplemental Information:

**Supplementary Table S1. Summary of Serious Adverse Events (SAEs) in the Non-MSKD Arm**

| Participant | Grade | Serious Adverse Event (SAE) |
|---|---|---|
| 01-003 | 3 | Hepatic infection |
| 01-006 | 3 | Hyperbilirubinaemia |
| 01-006 | 3 | Gastrointestinal haemorrhage |
| 01-020 | 2 | Pneumonia |
| 01-020 | 3 | Pleural effusion |
| 01-022 | 3 | Sepsis |
| 01-022 | 3 | Neutrophil count decreased |
| 01-022 | 3 | Anaemia |
| 01-022 | 3 | Haematuria |
| 01-022 | 3 | Platelet count decreased |
| 01-026 | 3 | Sepsis |
| 01-026 | 3 | Mucosal inflammation |
| 04-023 | 3 | Atrial flutter |
| 04-025 | 2 | Pyrexia |
| 04-025 | 3 | Hyperbilirubinaemia |
| 04-025 | 3 | Sepsis |
| 04-031 | 3 | Intra-abdominal haemorrhage |
| 04-031 | 3 | Gastrointestinal haemorrhage |
| 04-035 | 3 | Abdominal pain |
| 04-040 | 3 | Febrile neutropenia |
| 04-040 | 3 | Platelet count decreased |
| 04-040 | 3 | Colitis |
| 04-040 | 3 | Abdominal pain |

Complete list of 23 SAEs, by participant.
A total of 10 participants had one or more SAE.

**Supplementary Table S2. Summary of Serious Adverse Events (SAEs) in the MSKD Arm**

| Participant | Grade | Serious Adverse Event (SAE) |
|---|---|---|
| 01-001 | 3 | Pneumonia |
| 01-007 | 3 | Nausea |
| 01-007 | 3 | Dehydration |
| 01-007 | 3 | Vomiting |
| 01-021 | 3 | Biliary tract infection |
| 04-009 | 3 | Embolism |
| 04-009 | 3 | Asthenia |
| 04-043 | 3 | Skin infection |
| 04-043 | 3 | Embolism |
| 04-043 | 3 | Dyspnoea |
| 07-024 | 5 | Accidental death (motorcycle accident) |

Complete list of 11 SAEs, by participant.
A total of 6 participants had one or more SAE.

### Figure S1: CONSORT Diagram

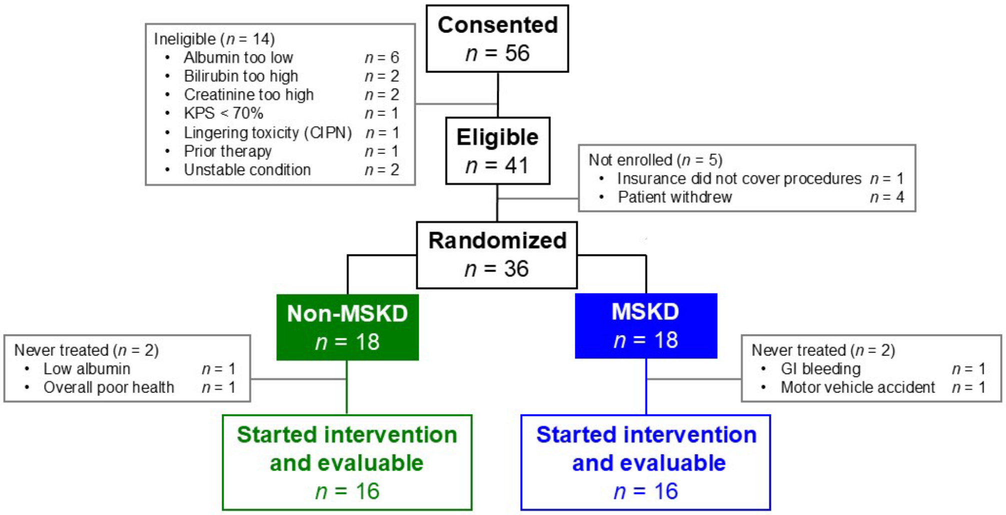
